# Molecular detection and phylogenetic characterization of hybrid *Fasciola* in black Bengal goats from Bangladesh: β-tubulin isotype 3 polymorphisms and genetic diversity

**DOI:** 10.1101/2025.06.12.25329487

**Authors:** Mohammad Manjurul Hasan, Nurnabi Ahmed, Babul Chandra Roy, Md. Rajiur Rahaman Rabbi, Md. Mahfuzur Rahman Sajib, Hiranmoy Biswas, Peru Gopal Biswas, Anisuzzaman, Mohammad Zahangir Alam, Md. Hasanuzzaman Talukder

## Abstract

Fasciolosis is a neglected tropical disease that poses a major threat to human and animal health, especially in low- and middle-income countries like Bangladesh, where climate change, widespread infection, emergence of hybrid *Fasciola* and rising benzimidazole (BZ) resistance have complicated disease control efforts. Therefore, this study aims to molecularly characterize and differentiate *F. gigantica* and hybrid *Fasciola* in black Bengal goats to better understand their genetic diversity and resistance patterns, ultimately supporting the development of more effective control strategies. A cross-sectional study was conducted from 2021–2022 across all eight divisions of Bangladesh, during which 3,134 liver flukes were collected from slaughterhouses. From these, 72 morphologically suspected hybrids were selected for molecular analysis. ITS2 sequencing and phylogeny confirmed all 72 as *F. gigantica*, revealing four haplotypes and low divergence from other Asian isolates. Hybrid status was confirmed in 76% of cases by *pepck* multiplex PCR and 39% by *pold* PCR-RFLP. Mitochondrial *NAD1* analysis of 25 hybrids revealed four haplotypes, moderate genetic diversity, and phylogenetic clustering with hybrids from India, China, Vietnam, and Nepal. *COX1* phylogeny supported these findings, placing Bangladeshi hybrids in a monophyletic *F. gigantica* clade, distinct from *F. hepatica*. Polymorphisms in the β-tubulin isotype 3 gene were observed in both *F. gigantica* and hybrid forms, with phylogenetic clustering alongside BZ-resistant *F. hepatica* isolates from Europe and Australia, suggesting possible emerging resistance in Bangladesh. This is the first report of hybrid *Fasciola* and potential resistance-associated β-tubulin variants in goats in Bangladesh. Whole genome sequencing and gene mapping related to pathogenicity and BZ resistance are imperative for future studies.

**Author summary:** Fasciolosis is a serious disease caused by parasitic worms that affects the health and productivity of both animals and people, particularly in poorer countries like Bangladesh. In our study, we focused on these parasites in native Black Bengal goats, which are an important source of income and nutrition for rural communities. We traveled across all eight divisions of Bangladesh and collected over 3,000 liver flukes from infected goats. We then used genetic tools to identify whether the parasites were a known species or a mix (hybrid) of two different types. We found that many of the parasites were hybrids, which are harder to detect and may resist common treatments. We also discovered genetic signs that these worms might be developing resistance to drugs commonly used to treat them. This is the first-time hybrid liver flukes have been reported in goats in Bangladesh. Our findings raise concern that current treatments may become less effective over time. We believe this work highlights the urgent need for better disease monitoring, improved treatment strategies, and further research using whole genome sequencing to understand how these parasites spread and survive.

## Introduction

Fasciolosis is a food-borne neglected tropical disease caused by infection with the liver flukes *Fasciola gigantica* and *F. hepatica*, negatively impacting global health and agriculture (1, 2, 3). This zoonotic helminth infection leads to an estimated loss of 2.4 million disability-adjusted life years (DALYs) annually across more than 75 countries with millions more at risk, particularly in low- and middle-income countries (3, 4). In livestock, the disease is even more severe, with approximately 700 million animals at risk, leading to substantial economic losses (5).

Climate changes and global warming have contributed to the increasing, incidence of fasciolosis, exacerbating the challenges of disease control. Resistance to broad-spectrum benzimidazole (BZ) anthelmintics, such as triclabendazole (TCBZ) and albendazole (ABZ), has emerged worldwide, complicating management strategies (6, 7). Our previous study evaluated the *in vitro* efficacy of nitroxinil (NTON), TCBZ, and ABZ against *F. gigantica* isolates by assessing motility, histopathological alterations, morphometric features, and ultrastructural changes using scanning electron microscopy. The findings revealed that NTON was the most potent, followed by TCBZ and ABZ, raising concerns about the development of TCBZ and ABZ resistance in *Fasciola* populations in Bangladesh (8).

The epidemiological landscape of fascioliasis has been further complicated by the emergence of hybrid, intermediate, and parthenogenetic *Fasciola* forms, which result from the co-infection of *F. gigantica* and *F. hepatica* within the same host (9). This phenomenon, particularly prevalent in Asia and Africa, (10, 11, 12, 13, 14, 15, 16) poses a significant threat to livestock production and human health by potentially increasing transmission rate and expanding the geographic distribution of the disease (17, 18). The urgency for sustainable control strategies, including vaccine development, rational anthelmintic use, improved farm management, and enhanced precision diagnostic tools, is paramount (19). Additionally, epidemiological surveillance of hybrid *Fasciola* and resistance patterns is crucial for implementing effective disease control policies.

Traditional methods for diagnosing and differentiating *Fasciola* species rely on morphological characteristics; however, precise identification is often challenging due to overlapping features (20). Molecular techniques have provided an alternative approach for species differentiation and population genetics analysis. Genetic markers, such as PCR-RFLP of the internal transcribed spacer (ITS1 & ITS2) regions (Anh et al., 2018; Mir et al., 2019; Saadatnia et al., 2022) and mitochondrial genes like cytochrome c oxidase (COX1) and NADH dehydrogenase subunit 1 (NAD1) (9, 12, 13) have proven valuable for distinguishing *F. gigantica* and *F. hepatica* (Calvani and Šlapeta, 2021). Moreover, recent studies have identified DNA polymerase delta (pold) and phosphoenolpyruvate carboxykinase (pepck) genes as novel genetic markers for discriminating *Fasciola* species, including hybrid forms (13, 21).

Tubulin proteins have been extensively studied in parasitic helminths due to their role as primary targets of benzimidazole drugs (22). Mutations in nematode tubulin genes can confer resistance to BZ treatments (6). Mutations in tubulin genes can confer resistance to BZ treatments, a phenomenon well-documented in nematodes such as *Haemonchus contortus*, where resistance arises from specific amino acid substitutions in β-tubulin isotype 1 (23). Recent studies have highlighted the β-tubulin 3 isotype as a promising molecular marker for detecting BZ-resistant *F. hepatica* (24, 25).

Bangladesh is a densely populated country with a staggering 200 million people and over 60 million ruminants is endemic to fascioliasis. The reported prevalence of fascioliasis in slaughtered animals varies widely, with infection rates of 15–66% in cattle, 3.8–22% in goats, 81% in sheep and 23–47% in buffaloes respectively (26, 27). However, the true burden, including subclinical infections, is likely much higher (26). The increasing development of BZ resistance, particularly in small ruminants, presents a growing threat to livestock production in Bangladesh (8). Additionally, the first reported presence of intermediate form of *Fasciola* in goats in Bangladesh (28) underscores the need for further investigation into its epidemiology, population structure, and genetic diversity.

Given the rising threat of anthelmintic resistance and the presence of hybrid *Fasciola* forms, it is crucial to generate comprehensive data on their genetic diversity and resistance status. Therefore, this study aims to precisely characterize and differentiate *F. gigantica* and hybrid/parthenogenetic *Fasciola* in goats in Bangladesh using well-established molecular markers. This research will provide valuable insights into their genetic makeup, epidemiology, disease transmission patterns, and resistance mechanisms, ultimately aiding policymakers in designing effective control strategies.

## Results

### Species confirmation by amplifying *ITS2* gene evolutionary divergence and phylogenetic characterization

A fragment of 508 bp of the *ITS2* gene was successfully amplified from all selected 72 *Fasciola* specimens supposed to be hybrid form from 3,134 flukes collected from slaughtered black Bengal goats in Bangladesh. After gel electrophoresis, all samples exhibited a distinct band corresponding to the expected 508 bp fragment. No non-specific bands were observed, indicating the specificity of the *ITS2* primers used in the study (Supplementary file: Figure S2).

For partial sanger sequencing, we randomly choose two positive PCR products from each division and a total of 15 ITS2 sequences were successfully produced and deposited GenBank under the accession number ON872490-ON8724504. The haplotype analysis of 15 sequences revealed the presence of four distinct haplotypes (h = 4), with a haplotype diversity (Hd) of 0.4667. The ITS2 nucleotide distribution analysis among haplotypes and reference sequences provides insights into the genetic variation within the dataset. Haplotype 1, haplotype 3, and haplotype 4 share an identical nucleotide composition with the KR5600070 *F. gigantica* India reference sequence, indicating a high degree of conservation. In contrast, haplotype 2 exhibits multiple nucleotide substitutions such as transitions (A ↔ G or C ↔ T) and transversions (A ↔ C, A ↔ T, G ↔ C, G ↔ T) at positions 101, 135, 139, 151, 175, 178, 184, 203, 204, 217, 220, 230, 234, 240, 252, 253, 283, 312, 316, 322, 328, 335, 337, 339, 344, 346, 358, 366, 408, 419, 425, 440, 443, 448, and 460 (Supplementary table 2), suggesting a distinct genetic profile. Additionally, the MK321644 hybrid *Fasciola* Chad sequence displays unique variations at positions 335, 346, 366, 397, 403, and 408, further indicating genetic divergence. The presence of a single dominant haplotype (Hap 1) in most samples suggests a conserved lineage, while the observed sequence divergence in Hap 2 and the hybrid sequence points to possible evolutionary pressures, geographical influences, or historical genetic recombination events shaping the genetic diversity within this population.

Estimates of evolutionary divergence between study origin ITS2 Sequences and best hit scoring ITS2 GenBank sequences were performed. Analyses were conducted using the Tamura-Nei model. This analysis involved 15 study generated *F. gigantica* ITS2 sequences and 10 *F. gigantica* ITS2 nucleotide sequences of India (KR560070), China (KF543340), Vietnam (MT429170), Kenya (MZ396907), Chad (MK321619), Cote d’voire (MK321627), Indonesia (PP693042), Zimbabwe (MW046876), Turkey (KY613944), Australia (MF678651) and one hybrid *Fasciola* ITS2 sequences from Chad (MK321644). All ambiguous positions were removed for each sequence pair (pairwise deletion option). There were a total of 1048 positions in the final dataset. Evolutionary analyses were conducted in MEGA11. The evolutionary divergence analysis of ITS2 sequences reveals a high degree of genetic similarity among the study samples (ON872490–ON872504), with pairwise distances ranging from 0.00 to 0.01. This suggests that the study-origin sequences belong to a closely related genetic cluster. When compared to GenBank reference sequences from different geographical regions, the study sequences exhibit minimal divergence (0.00–0.02) with *F. gigantica* (Fg) isolates from India, China, and Vietnam, indicating a possible shared ancestry or limited evolutionary separation. In contrast, moderate divergence (0.07–0.08) is observed with Fg isolates from Zimbabwe, Turkey, and Chad, suggesting greater genetic differentiation. These findings indicate potential geographic clustering, where the study sequences are more closely related to certain Asian isolates than to African or European counterparts. This data provides valuable insights into the genetic variation and potential phylogeographic distribution of the studied ITS2 sequences (Supplementary Table 3).

The phylogenetic tree has been constructed by using 55 ITS2 sequences where 15 sequences were study generated, 31 were best hit scoring *F. gigantica* sequences retrieved from NCBI gene bank and 8 were ITS2 sequences of *Fasciola hepatica* to compare. ITS2 sequence of *Paraphistomum leydeni* was used as an outgroup.

The tree revealed two distinct divided clades A and B (Figure 1). This neighbour joining tree demonstrated that all the study generated sequences formed a well-supported cluster within *F. gigantica*, closely related to isolates found in India, China, Indonesia, Vietnam, Egypt, Chad, Cote D’voire, Turkey, Zimbabwe and Kenya suggesting a common lineage. Notably, the phylogenetic tree distinguished *F. gigantica* from *F. hepatica*, a separate clade with high bootstrap support. These findings unveiled that different species of *Fasciola* has different genomic background and thus this species with same genomic background were clustered.

**Figure 1.**
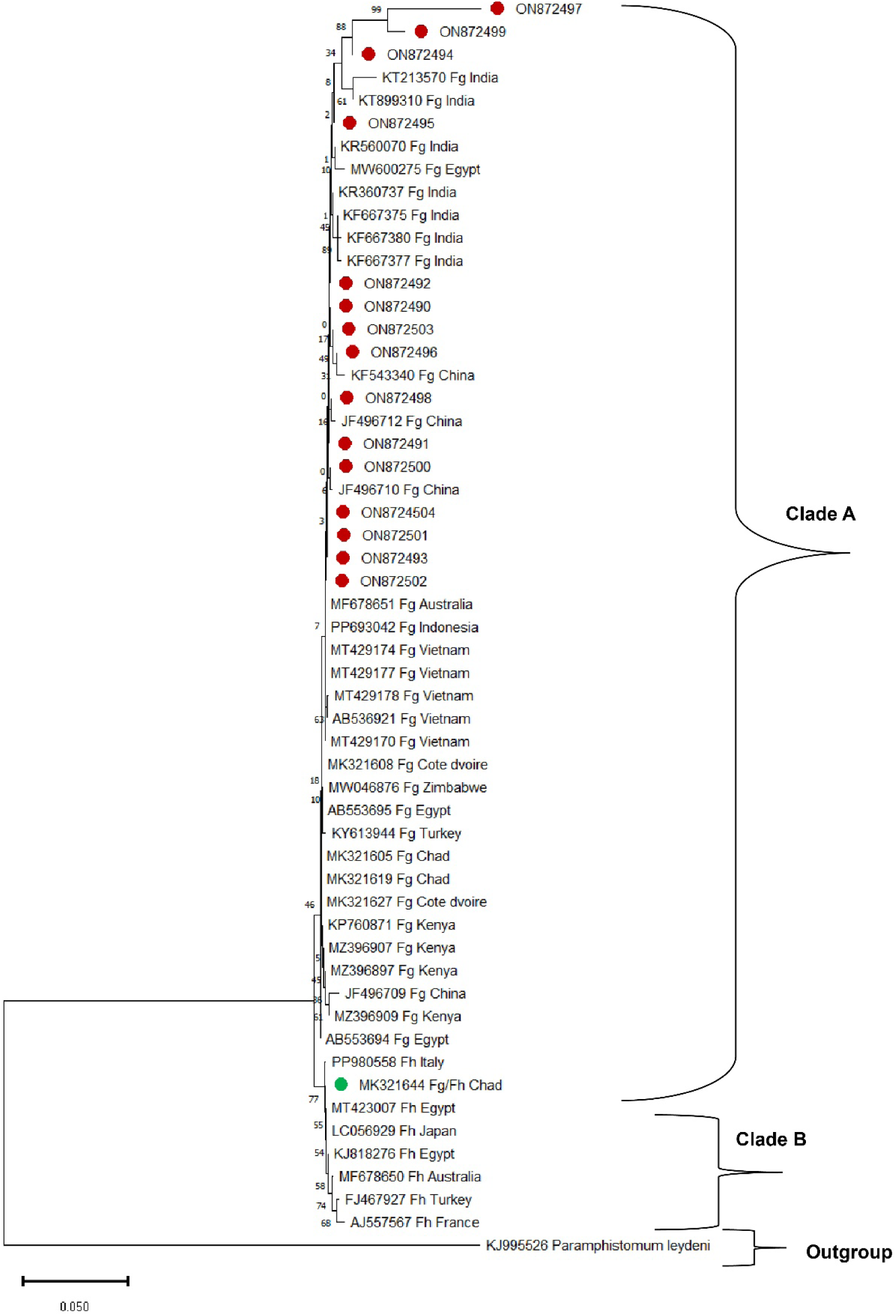
Neighbor-joining phylogenetic tree based on partial ITS2 sequences of Fasciola isolates from various hosts and geographical regions. *F. gigantica* (Fg) and *F. hepatica* (Fh) are represented, with *Paramphistomum leydeni* serving as the outgroup. Red-colored circle denotes study isolates. Green-colored circle denotes Fg/Fh isolate of Chad. The scale bar represents the proportion of nucleotide substitutions per site across the branches

### Molecular confirmation of hybrid form of *Fasciola* (Fg/Fh)

A total of 72 flukes genomic DNA were used to amplify two nuclear markers namely *Phosphoenolpyruvate carboxykinase* (*Pepck*) and *Polymerase delta* (*Pold*) through multiplex PCR and PCR-RFLP respectively. These two markers are the confirmatory markers for identifying hybrid *Fasciola* species. Species-specific bands for *Fasciola hepatica* (∼241 bp) and *Fasciola gigantica* (∼509 bp) were observed in 55 (76%) flukes from *Pepck* multiplex PCR products after gel electrophoresis that confirmed hybrid *Fasciola* species (Figure 2.A). Meanwhile 28 (39%) *Pold* PCR-RFLP product after digesting with *AluI* displayed distinct bands at 708 bp and 544 bp indicating *F. gigantica and F. hepatica* respectively (Figure 2.B).

**Figure 2.**
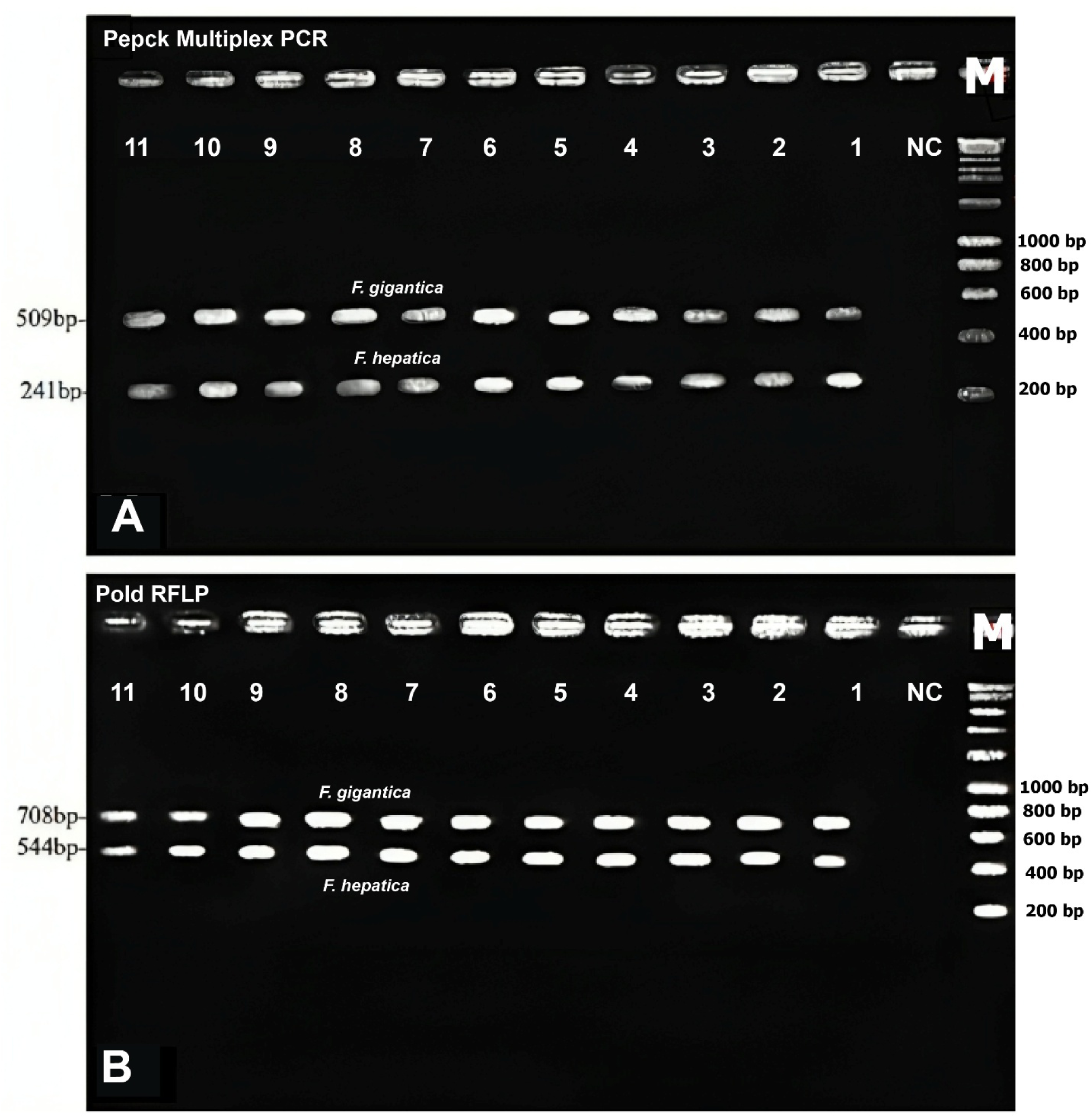
Representative gel image displaying: (A) *Pepck* multiplex PCR results. Our study samples exhibit species-specific bands at ∼249 bp and ∼510 bp, respectively. Hybrid *Fasciola* species show the presence of both bands. (B) *Pold* PCR-RFLP analysis. *Pold* PCR products were digested with *AluI*, yielding distinct banding patterns: *F. hepatica* at 708 bp and *F. gigantica* at 544 bp. Hybrid flukes exhibit a mixed fragment pattern, displaying bands at 708 and 544 bp respectively.

### Genotyping, haplotype analysis, nucleotide distribution and phylogenetic characterization of hybrid *Fasciola*

A fragment of 535 bp of the NAD1 gene and 430 bp of COX1 gene was successfully amplified by conventional PCR from 28 identified hybrid *Fasciola by Pepck* multiplex PCR and *Pold* PCR-RFLP (Figure 3).

**Figure 3:**
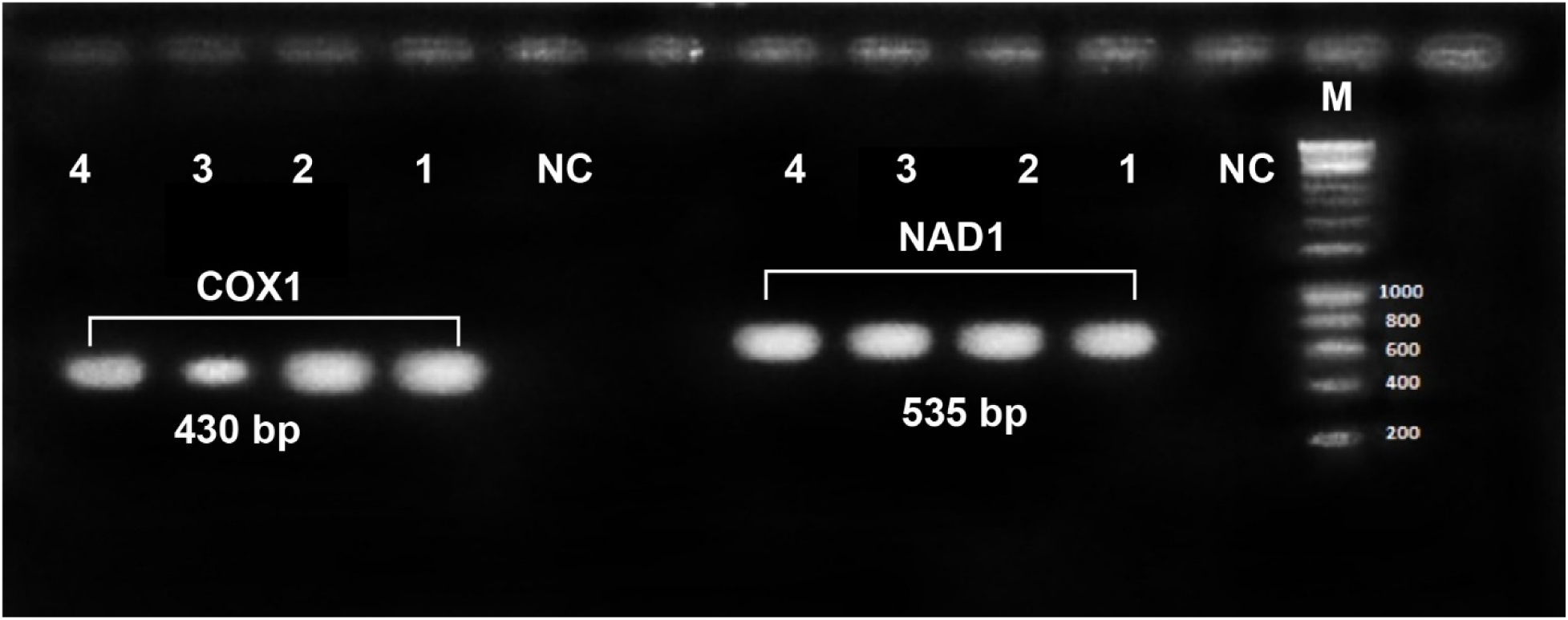
Image showing specific PCR amplifications of COX1 sequence (438 bp) and NADI sequence (535 bp) from hybrid Fg/Fh on 1.5% agarose gel [Lane M (Marker-100 bp)]

Twenty-five DNA sequences were produced successfully from 28 NAD1 gene positive amplicons while 23 DNA sequences were produced successfully from 28 COX1 gene positive amplicons (Sequences are in the supplementary file).

We only described genotyping, haplotype analysis and nucleotide distribution for NAD1 gene as both were mitochondrial gene. The haplotype analysis of 25 NAD1 sequences, covering a 446 bp region without alignment gaps, showed 12 parsimony informative sites (positions 132, 386, 387, 388, 389, 390, 391, 392, 393, 403, 404, 405) yielding 4 haplotypes with haplotype diversity 0.7267. Among them, haplotype 2 was the most frequent, comprising 10 sequences from multiple locations, including Mymensingh (Ms), Chattogram (Ctg), Rajshahi (Rs), and Khulna (Kl). Haplotype 4 was the second most prevalent, consisting of 8 sequences predominantly from Barishal (Bs) and Rangpur (Rp). Haplotype 1 and Haplotype 3 were less common, containing 4 and 3 sequences, respectively (Details are given in Supplementary file). The geographic distribution of haplotypes suggests a moderate level of genetic variation, with some haplotypes shared across multiple regions, while others are more localized. The presence of 12 variable sites indicates a degree of polymorphism in the dataset, contributing to the observed haplotype diversity.

The pairwise genetic distance analysis of partial NAD1 sequences (Table 2) compares the genetic divergence between study-generated hybrid *Fasciola* (Fg/Fh) haplotypes and the best hit-scoring Fg/Fh isolates from different countries retrieved from GenBank. Genetic distances range from 0.00 to 0.06, indicating varying levels of similarity among the isolates. Among the study-generated haplotypes, BD-NAD1-Fg/Fh-Haplotype-01, -02, -03, and -04 exhibit low genetic distances (0.00–0.03), suggesting high similarity. Haplotype-01 shows no divergence (0.00) from the Bangladeshi Fg/Fh isolate (AB894372) and other GenBank sequences from India (LC012896.1), Nepal (AB894348.1), China (AB604941.1, AB477368.1), and Vietnam (AB536764.1), indicating close genetic relationships. Haplotype-02 has slightly higher divergence (0.02–0.03) from these isolates, while haplotype-03 maintains 0.01–0.02 genetic distance. Notably, the Egyptian isolate (LC076202.1) exhibits the highest genetic distance (0.04– 0.06) from the Bangladeshi haplotypes and other Asian isolates, suggesting greater genetic divergence. The Vietnamese isolate (AB536764.1) also shows slightly higher distances (0.01) compared to other Asian isolates. The nucleotide distribution analysis shows genetic differences among hybrid *Fasciola* (Fg/Fh) haplotypes and reference isolates from GenBank. BD-NAD1-Fg/Fh-Haplotype-01 is identical to the reference isolates at the examined positions, while haplotypes 02 and 03 have several nucleotide changes, particularly at positions 426, 428, 429, 430, 431, 432, and 445. Haplotype-04 is similar to Indian and Nepalese isolates but differs at position 427. These changes include six transitions (A↔G or C↔T) and eight transversions (A↔T or G↔C), with a Ts/Tv ratio of 0.759 (Table 3).

**Table 1.**
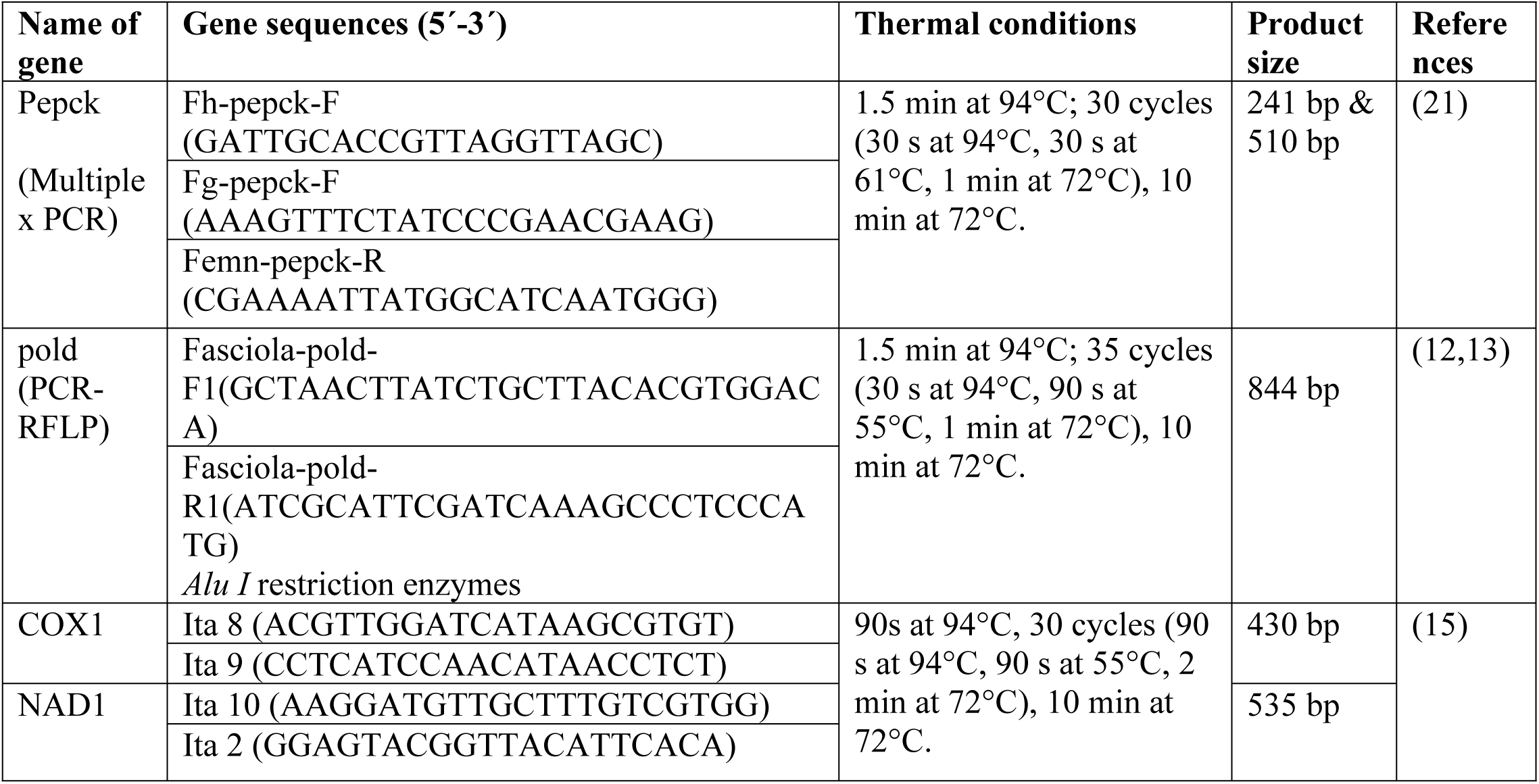
Primer sets used in the study and their thermal conditions to detect hybrid *Fasciola* and their phylogenetic characterization.

**Table 2.** Pairwise genetic distance analysis of partial NAD1 sequences between study-generated hybrid *Fasciola* (Fg/Fh) isolates and best hit-scoring Fg/Fh isolates from various countries retrieved from GenBank.

| Sequences | 1 | 2 | 3 | 4 | 5 | 6 | 7 | 8 | 9 | 10 | 11 |
| --- | --- | --- | --- | --- | --- | --- | --- | --- | --- | --- | --- |
| BD-NAD1-Fg/Fh-Haplotype-01 | - |  |  |  |  |  |  |  |  |  |  |
| BD-NAD1-Fg/Fh-Haplotype-02 | 0.02 | - |  |  |  |  |  |  |  |  |  |
| BD-NAD1-Fg/Fh-Haplotype-03 | 0.01 | 0.01 | - |  |  |  |  |  |  |  |  |
| BD-NAD1-Fg/Fh-Haplotype-04 | 0.00 | 0.03 | 0.02 | - |  |  |  |  |  |  |  |
| AB894372 Fg/Fh goat Bangladesh | 0.00 | 0.02 | 0.01 | 0.00 | - |  |  |  |  |  |  |
| LC012896.1 Fg/Fh Cattle India | 0.00 | 0.02 | 0.01 | 0.00 | 0.00 | - |  |  |  |  |  |
| AB894348.1 Fg/Fh Cattle Nepal | 0.00 | 0.02 | 0.01 | 0.00 | 0.00 | 0.00 | - |  |  |  |  |
| AB604941.1 Fg/Fh Cattle China | 0.00 | 0.02 | 0.01 | 0.00 | 0.00 | 0.00 | 0.00 | - |  |  |  |
| AB536764.1 Fg/Fh Cattle Vietnam | 0.01 | 0.02 | 0.01 | 0.01 | 0.01 | 0.01 | 0.01 | 0.01 | - |  |  |
| LC076202.1 Fg/Fh Sheep Egypt | 0.04 | 0.06 | 0.05 | 0.04 | 0.04 | 0.04 | 0.04 | 0.04 | 0.05 | - |  |

**Table 3.** Nucleotide distribution of study generated partial NAD1 gene sequences of hybrid *Fasciola* with reference sequences retrieved from GenBank.

| Sequences | Nucleotide distribution of partial NAD1 gene |  |  |  |  |  |  |  |  |  |  |  |  |  |  |  |  |  |  |  |  |  |  |  |  |  |  |
| --- | --- | --- | --- | --- | --- | --- | --- | --- | --- | --- | --- | --- | --- | --- | --- | --- | --- | --- | --- | --- | --- | --- | --- | --- | --- | --- | --- |
|  | 81 | 99 | 117 | 159 | 171 | 196 | 207 | 282 | 285 | 303 | 355 | 381 | 409 | 453 | 460 | 508 | 513 | 514 | 528 | 529 | 530 | 538 | 551 | 567 | 568 | 573 | 574 |
| AB894372.1 Fg/Fh Bangladesh | G | G | T | G | T | C | A | A | G | G | A | G | C | G | G | G | C | - | G | T | T | A | T | G | G | - | T |
| Bangladesh-Fg/Fh-Hap1 | . | . | . | . | . | . | . | . | . | . | . | . | . | . | . |  | T | A | . | . | . | . | . | . | . | G | . |
| Bangladesh-Fg/Fh-Hap2 | . | . | . | . | . | . | . | . | . | . | . | . | . | . | . | . | . | - | . | . | . | . | . | . | . | - | . |
| LC012896.1 Fg/Fh Cattle India | . | . | . | . | . | . | . | . | . | . | . | . | . | . | . |  | T | - | . | . | . | . | . | . | . | - | . |
| AB894348.1 Fg/Fh Cattle Nepal | . | . | . | . | . | . | . | . | . | . | . | . | . | . | . | . | . | - | . | . | . | . | . | . | . | - | . |
| AB604941.1 Fg/Fh Cattle China | . | . | . | . | . | . | . | . | . | . | . | . | . | . | T | . | . | - | . | . | . | G | . | . | . | - | . |
| AB536764.1 Fg/Fh Cattle Vietnam | . | . | . | . | . | . | . | . | . | . | . | . | . | . | . | . | . | - | T | C | G | . | . | . | . | T | . |
| LC076202.1 Fg/Fh Sheep Egypt | T | A | C | A | C | T | T | G | T | A | G | A | T | A |  | A | . | - | . | . | . | . | C | T | T | - | C |

Phylogenetic tree was built with all study generate NAD1 (Figure 4) and COX1 sequences (Figure 5) separately with best hit-scoring NAD1 and COX1 sequences of hybrid *Fasciola*, *F. gigantica* and *F. hepatica* isolates across the world retrieved from GenBank. Neighbour-Joining (NJ) and Maximum Likelihood (ML) were used to produce phylogenetic tree that illustrate same findings in case of both phylogenies. Readers better understanding, only the neighbour-joining trees have been incorporated, visualized and explained here. The Neighbour-Joining (NJ) phylogenetic tree for the NAD1 gene, constructed with 1000 bootstrap replicates, revealed two major clades (A and B), with clade A further subdivided into multiple subclades. In subclade I of clade A, Fg/Fh isolates from Bangladesh (BD-Ms, BD-SL, BD-Rs, BD-Rp, BD-Dh, BD-KL, BD-Ctg, and BD-Bs) clustered closely with Fg/Fh isolates from India (LC012896.1), China (AB604941.1, AB604942.1, AB477368.1), Vietnam (AB536764.1), Nepal (AB894348.1) while study Fg/Fh isolates also clustered closely with *Fasciola gigantica* (Fg) isolates from these above mentioned country and Iran (KX036360), which is supported by high bootstrap values (>75%). This clustering pattern suggests a strong genetic similarity among these geographically diverse isolates, indicating potential regional transmission dynamics or a shared evolutionary origin. Additionally, a distinct subclade included *Fasciola* gigantica isolates from sheep in Armenia (MG972404) and India (ON641029), further supporting genetic divergence within the *Fasciola* population. Interestingly, no Fg/Fh isolates of our study form cluster with *Fasciola hepatica* isolates from temperate region suggesting further genetic analysis is needed to determine whether multiple hybridization and introgression events have occurred within this region.

**Figure 4.**
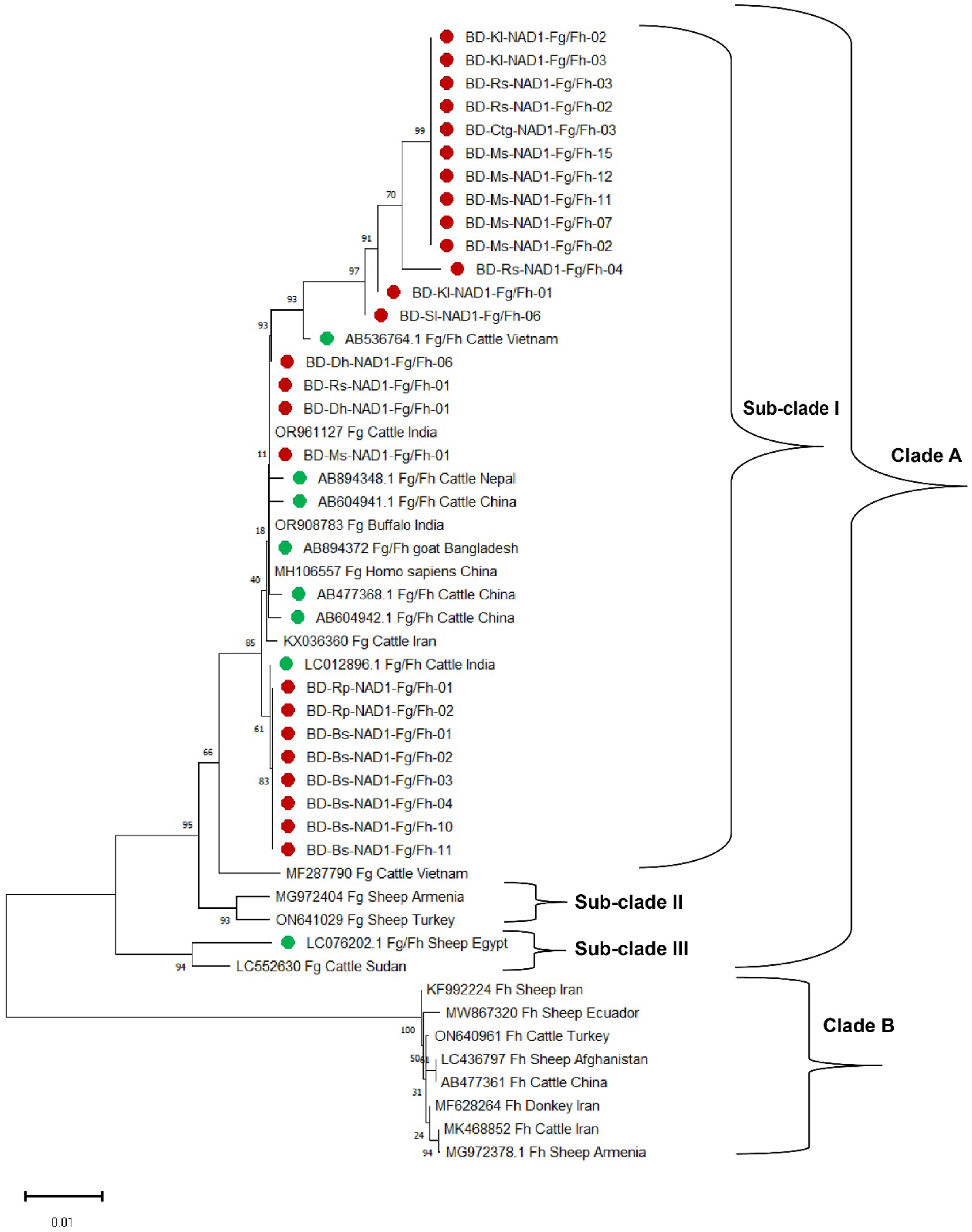
Neighbor-joining phylogenetic tree based on partial NAD1 gene sequences of Fg/Fh isolates from various hosts and geographical regions. *F. gigantica* (Fg) and *F. hepatica* (Fh) are represented. Red-colored circle denotes study isolates. Green-colored circle denotes Fg/Fh isolates of different countries. The scale bar represents the proportion of nucleotide substitutions per site across the branches

**Figure 5.**
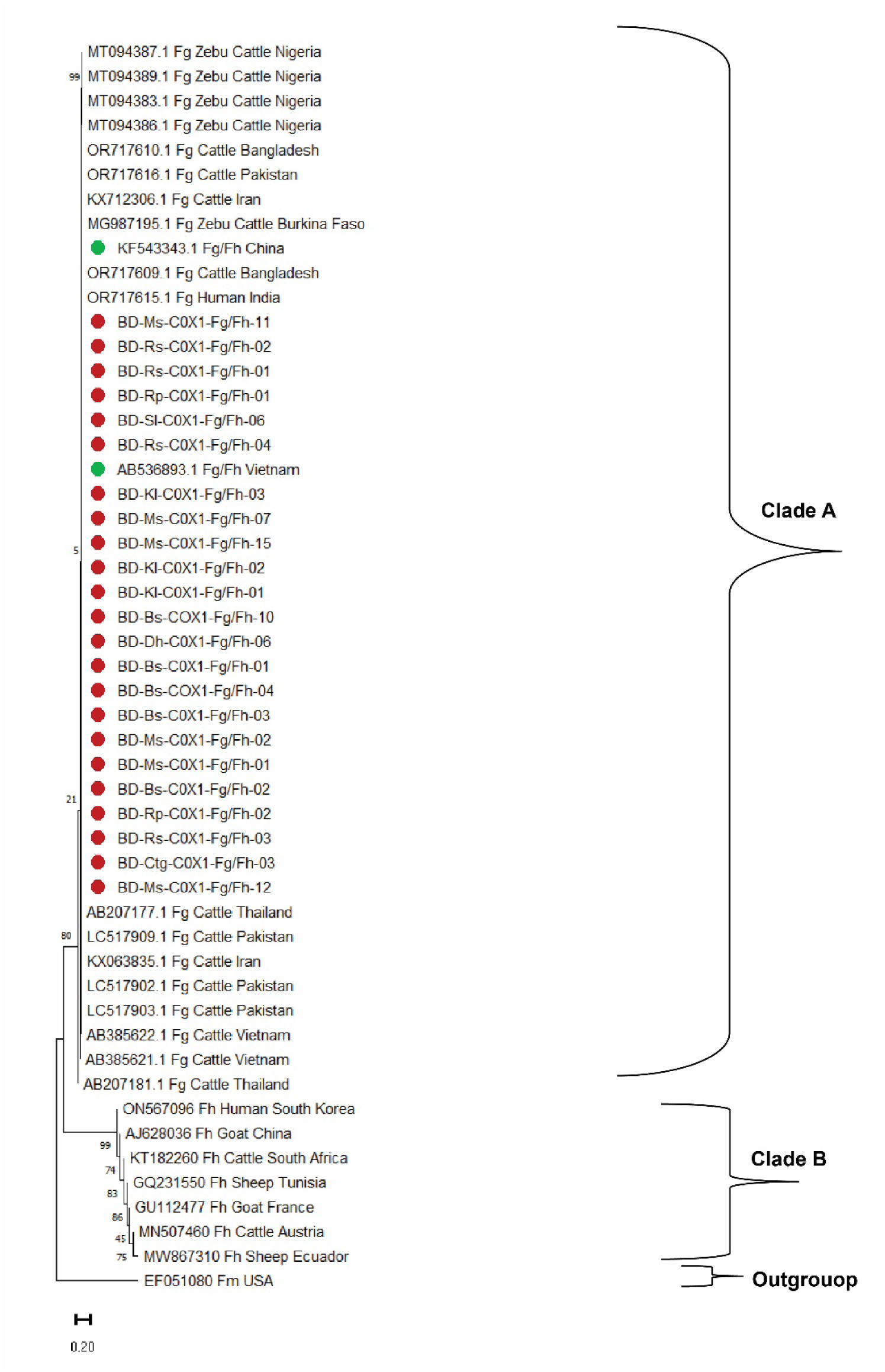
Neighbor-joining phylogenetic tree based on partial COX1 gene sequences of Fg/Fh isolates from various hosts and geographical regions. *F. gigantica* (Fg) and *F. hepatica* (Fh) are represented. Red-colored circle denotes study isolates. Green-colored circle denotes Fg/Fh isolates of different countries. The scale bar represents the proportion of nucleotide substitutions per site across the branches

The COX1 gene tree delineates two major clades, Clade A and Clade B, with strong bootstrap support (Figure 5). Clade A comprises *Fasciola gigantica* and hybrid *Fasciola* (Fg/Fh) isolates from multiple geographic locations, including Bangladesh, Nigeria, China, Vietnam, Pakistan, India, Iran, and Thailand. Within Clade A, hybrid *Fasciola* (Fg/Fh) isolates from Bangladesh (BD) formed a well-supported monophyletic group, clustering closely with Fg/Fh isolates from Vietnam (AB536893.1) and China (KF543343.1). Furthermore, study isolates exhibited no clear geographic sub-structuring and were interspersed with other *F. gigantica* or Fg/Fh sequences from different countries, suggesting genetic similarity across regions.

On the other hand, Clade B primarily consists of *Fasciola hepatica* isolates, clustering separately from *F. gigantica* with a high bootstrap value of 99%, confirming species-level differentiation. This clade includes isolates from South Korea, China, France, South Africa, Austria, and Tunisia.

### Performing Polymerase Chain Reaction (PCR) for β-tubulin gene isotype 3 polymorphisms of *F. gigantica* and hybrid form (Fg/Fh)

β-tubulin isotype 3 gene were amplified from 24 *F. gigantica* (three from each division were randomly selected) and 55 identified Fg/Fh isolates through conventional PCR. At least one *F. gigantica* from each division and 29 Fg/Fh isolates showed band of 935 bp (Figure 6). This β-tubulin isotype 3 gene polymorphism in *F. gigantica* and Fg/Fh isolates from goat in Bangladesh have been identified for the first time.

**Figure 6.**
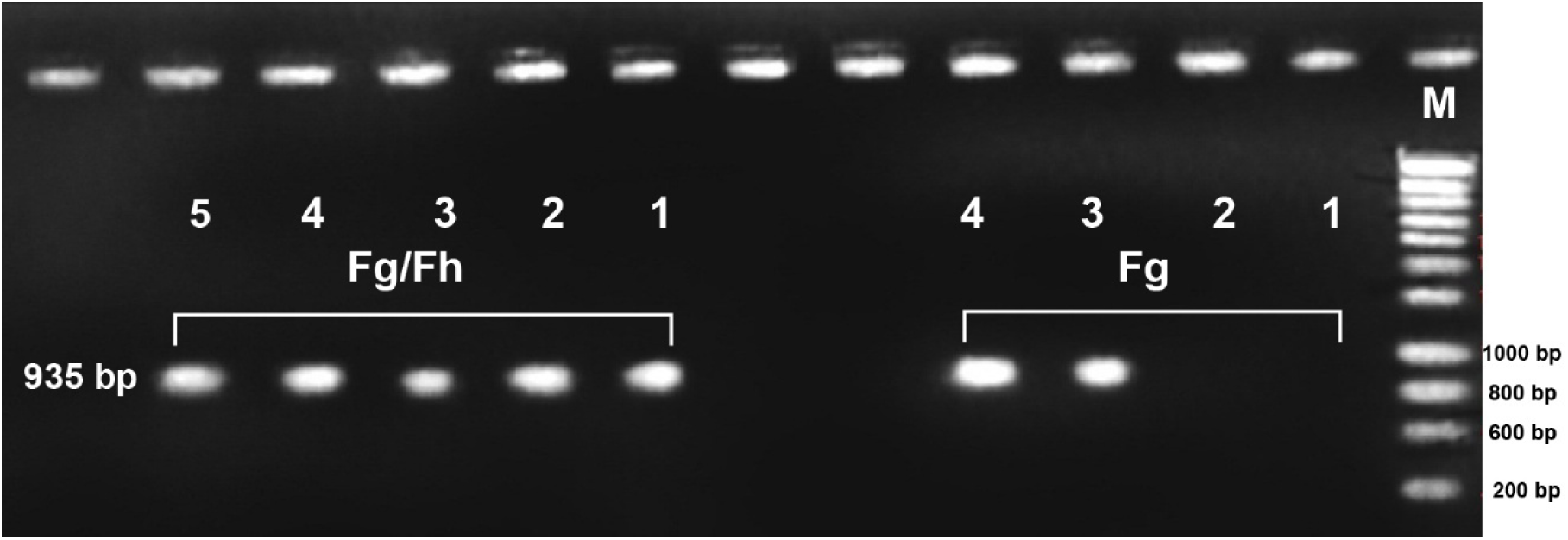
Representative gel image displaying conventional PCR results. DNA samples of *F. gigantica* and hybrid form (Fg/Fh) exhibit species-specific bands at ∼935 bp. M: denotes DNA ladder

### Phylogenetic characterization of Beta tubulin isotype 3 polymorphism in *F. gigantica* and hybrid *Fasciola* (Fg/Fh)

Neighbour-Joining (NJ) and Maximum Likelihood (ML) methods were employed to construct the phylogenetic tree based on the β-tubulin isotype 3 (bt3) gene, both producing congruent topologies (Figure 7). The NJ phylogenetic tree, generated using 1000 bootstrap replicates, delineates two primary clades (A and B), each reflecting distinct evolutionary relationships among *Fasciola* isolates. Within clade A, our study generated partial sequences of β-tubulin isotype 3 (bt3) gene from hybrid *Fasciola* form a well-supported cluster (bootstrap >90%) with benzimidazole resistant *Fasciola hepatica* isolates from Greece (HM535870, HM535875, HM535929, HM535937, HM535947), the United Kingdom (AM77765, AM933587), and Australia (MK248761, MK248816, MK248794) suggesting strong genetic relatedness among these geographically distant populations. However, our study generated all *Fasciola gigantica* isolates along with other hybrid *Fasciola* species distributed across multiple subclades within Clade B and form a distinct lineage. The presence of Bangladeshi isolates within different subclades suggests genetic heterogeneity, possibly arising from regional selection pressures, host specificity, or restricted gene flow. This study provides the first phylogenetic insight into bt3 gene diversity among *Fasciola* species in Bangladesh.

**Figure 7:**
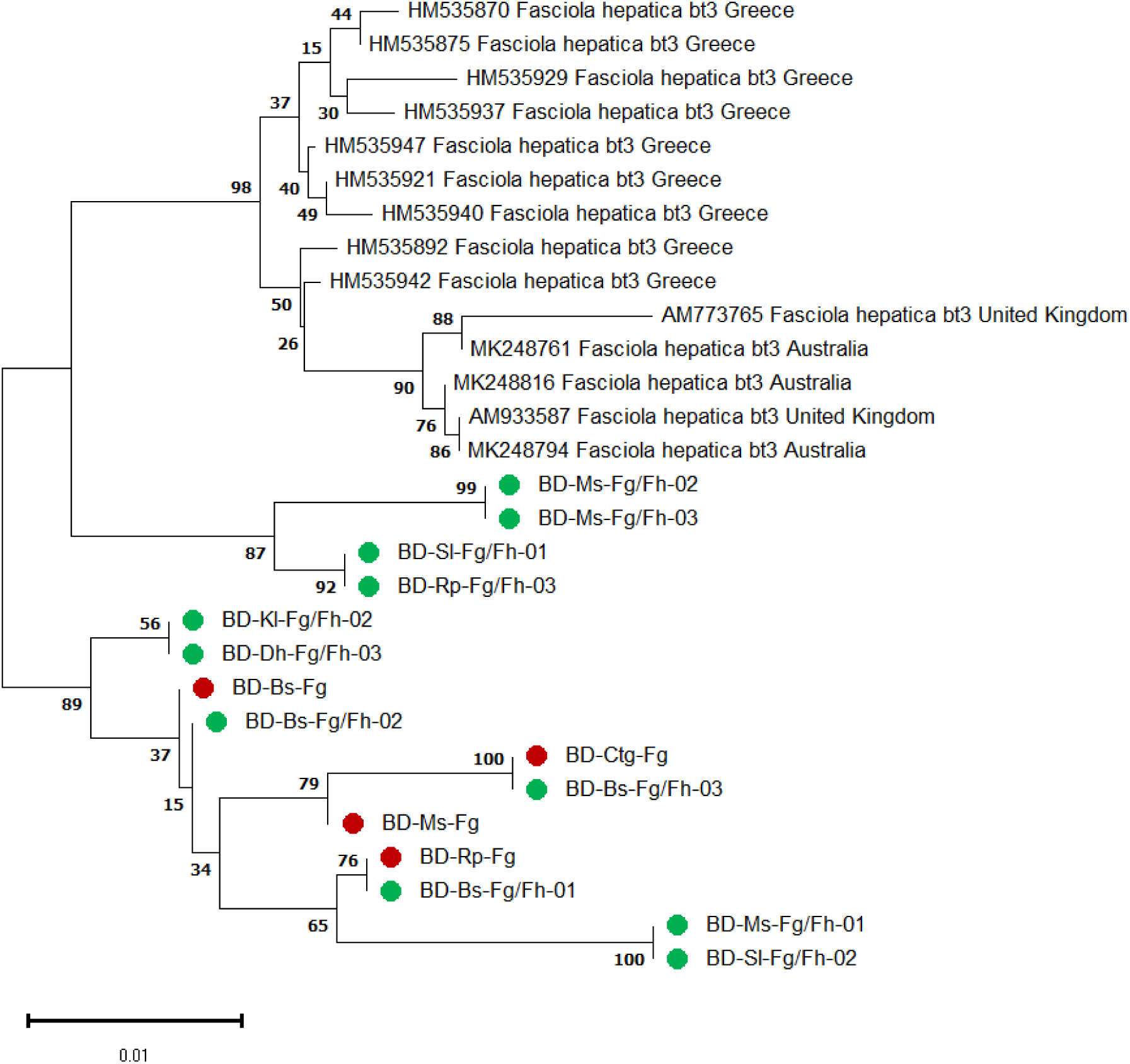
Neighbour-Joining (NJ) phylogenetic tree based on the β-tubulin isotype 3 (*bt3*) gene, constructed using 1000 bootstrap replicates. The tree illustrates the evolutionary relationships among *Fasciola* isolates (*F. gigantica*-red circled and Hybrid *Fasciola*-Green circled) from Bangladesh (BD) and various best hit-scoring isolates retrieved from GenBank. Bootstrap values are indicated at the nodes, representing statistical support for each branch.

## Discussions

This study provides the first comprehensive molecular evidence of hybrid *Fasciola* and potential benzimidazole (BZ) resistance in black Bengal goats in Bangladesh. Our molecular characterization revealed that all 72 morphologically suspected hybrid flukes were genetically identified as *F. gigantica* via ITS2 sequencing and phylogenetic analysis, forming a distinct cluster with other Asian *F. gigantica* isolates. The ITS2 region exhibited four haplotypes, suggesting some level of intra-species diversity. The low evolutionary divergence from other Asian isolates indicates a shared genetic background, possibly due to historical livestock movement or cross-border transmission. Our findings of ITS2-based identification of *F. gigantica* and four distinct haplotypes are consistent with previous research from India, Egypt, and Iran, where ITS2 proved effective for species delimitation in *Fasciola* spp. (29, 30)

To confirm hybrid status, we used two nuclear markers. The pepck multiplex PCR assay identified 76% of isolates as hybrids, whereas pold PCR-RFLP confirmed hybrid status in 39%. The present results were consistent with the findings reported earlier from Asian countries where the authors found 13.2% -90.9% *F. gigantica* and 9.1% - 63.0% parthenogenic *Fasciola,* respectively (9, 12, 13, 14, 21, 31, 32). The variation between two molecular techniques variation likely reflects differences in the sensitivity and resolution of these assays, as previously observed in studies from Nepal (31) and Vietnam (33) suggesting a widespread presence of hybrid *Fasciola* in South Asia, especially in areas where the geographic distribution of *F. hepatica* and *F. gigantica* may overlap due to climate shifts or animal movement (9). The high hybrid detection rate with *pepck* suggests its robustness as a diagnostic marker in identifying cryptic hybridization events (12).

Mitochondrial NAD1 analysis of 25 confirmed hybrids revealed four haplotypes and moderate genetic diversity (haplotype diversity = 0.7267), consistent with prior reports from India, China, and Nepal (32, 34). Our mitochondrial NAD1 and COX1 phylogenies showed strong relationships with other Asian hybrid isolates, supporting the hypothesis of regional gene flow and evolutionary conservation of hybrid forms. COX1 phylogeny corroborated these results, placing Bangladeshi hybrids within a monophyletic *F. gigantica* clade, distinct from *F. hepatica*, similar to the findings of Amor et al. (2011) (35). However, the presence of nuclear hybrid genotypes implies that historical hybridization events still leave a genomic signature in local populations.

Importantly, β-tubulin isotype 3 gene sequencing revealed polymorphisms in both *F. gigantica* and hybrid isolates. The phylogenetic clustering of Bangladeshi hybrids with BZ-resistant *F. hepatica* strains from Europe and Australia is alarming and suggests the potential emergence of drug-resistant genotypes in Bangladesh. Studies in the UK and Australia (36, 37) reported specific SNPs (e.g., F200Y, E198A) associated with BZ resistance. While we did not confirm these specific mutations, the phylogenetic proximity hints at the convergent evolution of resistance traits.

The confirmation of hybrid *Fasciola* with moderate genetic diversity and potential BZ resistance in black Bengal goats has significant implications for disease control in Bangladesh. The presence of hybrids may alter pathogenicity, host specificity, transmission dynamics, and drug response profiles, making them harder to manage using conventional approaches. Moreover, goats—often overlooked in national parasite control strategies—could act as reservoir hosts, sustaining hybrid transmission cycles and spreading resistant genotypes across livestock populations. The detection of resistance-associated β-tubulin polymorphisms warrants immediate attention. Continued use of BZ drugs such as TCBZ without monitoring for efficacy may accelerate resistance development, undermining treatment programs and increasing disease burden. Integrated parasite management, routine resistance surveillance, and diversification of treatment options are urgently needed. The clustering of hybrid isolates with known BZ-resistant strains from other continents suggests a potential early-stage resistance scenario in Bangladesh. If unchecked, this could lead to the spread of resistance alleles, especially under selective pressure from overuse or improper dosing of BZ drugs. Since β-tubulin is a target of BZs, mutations in this gene could reduce drug binding efficiency, as seen in resistant *F. hepatica* (6). These findings underline the importance of functional validation of β-tubulin mutations and the need for alternative diagnostic markers to detect early resistance. Whole-genome and transcriptomic studies could help identify other resistance-associated loci and provide a better understanding of the mechanisms involved.

## Limitations and Future directions

Our study had some limitations. Firstly, sample size for mitochondrial and β-tubulin analyses was relatively small compared to the total fluke population. This may have limited the detection of additional rare haplotypes or novel polymorphisms. Secondly, we did not perform functional validation of resistance mutations, making it difficult to confirm their phenotypic effects. Finally, environmental and host factors influencing hybridization and resistance were not addressed.

Future studies should focus on whole genome sequencing (WGS) and gene mapping to investigate the genomic architecture of hybrid *Fasciola*, especially in relation to drug resistance and pathogenicity. Functional assays such as BZ binding studies, gene editing, or expression analysis of resistance markers are essential to confirm the role of observed polymorphisms. Additionally, surveillance programs incorporating goats and other small ruminants should be expanded to monitor hybrid transmission and resistance trends across Bangladesh.

## METHODS

### Ethical statement

The study protocol was reviewed and approved by the Animal Welfare and Experimentation Ethics Committee (AWEEC) of Bangladesh Agricultural University, Mymensingh [Study No. AWEEC/BAU/2019(2)]. All procedures adhered to the highest ethical standards as per institutional guidelines.

### Study design and sampling procedure

This cross-sectional study was conducted between 2021 to 2022 in different local slaughterhouses of eight divisional cities of Bangladesh. The divisional cites are Dhaka, Rajshahi, Mymensingh, Chattogram (previously known as Chittagong), Khulna, Barishal, Sylhet and Rangpur (Supplementary file: Figure S1). These cities represent distinct agro-ecological zones of the country (38). The largest slaughterhouse located in each city was selected purposively. Sampling occurred monthly over the study period, following written consent from slaughterhouse authorities. Up to five infected livers and bile ducts were collected per day from each slaughterhouse during post-mortem inspection. In total, 376 *Fasciola*-infected livers and bile ducts were collected and transported under cold chain conditions to the diagnostic parasitology laboratory of the Department of Parasitology, Bangladesh Agricultural University, for further processing.

### Parasitological investigations

Livers and bile ducts were dissected into small cubes, and flukes were carefully separated using forceps and placed in 0.9% saline. A total of 3,134 flukes were recovered, with an average of eight per liver, ranging from a minimum of three to a maximum of 76. Gross examination identified 72 potential hybrid *Fasciola* specimens (Supplementary Table 1) as described previously which were later stored at -20°C for molecular analysis (13).

### DNA extraction from suspected hybrid liver flukes

A ∼20 mg section from the anterior part of each suspected hybrid fluke was excised and placed in a 1.5 ml eppendorf tube. Genomic DNA was extracted using the Monarch Genomic DNA Purification Kit (New England Biolabs, USA) according to the manufacturer’s instructions. DNA concentration was quantified using a NanoDrop spectrophotometer (Thermo Scientific, USA) at the department of pathology, faculty of Veterinary Science, and stored at -20°C until further analysis.

### Genotypic characterization and species identification of *Fasciola* spp

To confirm *Fasciola gigantica*, ribosomal rDNA ITS2 gene was amplified via conventional PCR following previously described protocol (39) from all 72 flukes genomic DNA (Supplementary Table 1). Each 25 µl reaction contained 1 µl of 10 pmol ITS2-F (TGTGTCGATGAAGAGCGCAG) and ITS2-R (TGGTTAGTTTCTTTTCCTCCGC) primers, 12.5 µl Takara master mix (Takara Bio USA, Inc.), 2.5 µl genomic DNA, and 8 µl deionized water. PCR cycling conditions included an initial denaturation at 94°C for 90 s, followed by 30 cycles of 94°C for 90 s, 55°C for 90 s, and 72°C for 120 s, with a final extension at 72°C for 10 min (39). The amplified PCR products were visualized using 1.8% agarose gels staining with ethidium bromide (40). From each division, two best band showing PCR positive amplicons were sequenced using an Applied Biosystems 3730XL automated DNA sequencer. Sequences were edited and trimmed using BioEdit software and the resultant sequences were aligned using ClustalW in MEGA11 with a gap opening penalty of 15.00 and a gap extension penalty of 6.66 (Nehra et al., 2022). The haplotypes were identified by DnaSP (Version 6.12.03). The DNA sequences were then compared with best hit-scoring sequences retrieved from GenBank representing *Fasciola* spp. across the world. Phylogenetic trees were constructed using the neighbor-joining method with the Tamura-Nei evolutionary model, selected based on the lowest Bayesian Information Criterion (BIC) and Akaike Information Criterion corrected (AICc) values in MEGA11 (41).

### Molecular Identification of Hybrid/Parthenogenetic/intermediate form *Fasciola*

Two nuclear markers, phosphoenolpyruvate carboxykinase (Pepck) multiplex PCR and polymerase delta (Pold) PCR-Restriction Fragment Length Polymorphism (PCR-RFLP), were employed to identify hybrid *Fasciola* as described previously (21). Primer sequences and thermal conditions used in two molecular techniques are detailed in Table 1. For Pepck multiplex PCR, each 25 µl reaction included 12.5 µl Takara master mix, 1 µl of each primer (10 pmol), 2.5 µl DNA, and 7 µl deionized water. PCR products were electrophoresed on 2% agarose gels stained with ethidium bromide. PCR-RFLP reactions followed the same protocol, with an additional digestion step using Alu I restriction enzyme (Toyobo, Osaka, Japan) at 37°C for 3 hours before visualization on 2% agarose gel.

### Diversity indices and Phylogenetic characterization of hybrid *Fasciola*

The fragments of NAD1 (535bp) and COX1 (430bp) genes were amplified through a PCR assay previously described previously (15). PCR reactions were performed under standard conditions, and four PCR amplicons per division were sequenced using an Applied Biosystems 3730XL automated DNA sequencer. Sequences were edited, trimmed and aligned following same protocol describe in this manuscript elsewhere. Diversity indices such as haplotype diversity, nucleotide diversity, nucleotide distribution was computed only for NAD1 gene as both NAD1 and COX1 were mitochondrial gene while phylogenetic characterization was done for both gene. Haplotype diversity and pairwise comparisons were evaluated using Mega 11.0 (42). The population diversity indices, including the number of haplotypes, haplotype diversity, and nucleotide diversity evaluated using DnaSP version 5.1 (43). Phylogenetic trees for NAD1 and COX1 gene were built by comparing study generated sequences with best hit scoring reference sequences from GenBank representing *Fasciola* isolates from South Asia, East and Southeast Asia, the Middle East, and the Americas via BLAST as described elsewhere in the manuscript.

### Molecular detection and phylogenetic characterization of beta-tubulin isotype 3 in *Fasciola gigantica* and hybrid *Fasciola*

A 935 bp fragment of the β-tubulin isotype 3 gene was amplified from randomly selected 15 *Fasciola gigantica* and hybrid *Fasciola* using primers β-tub295f (5’-AAYAAYTGGGCYAARGGNCAYTA-3’) and β-tub1230r (5’-TCRGTRAAYTCCATYTCRTCCAT-3’), followed by sequencing with an Applied Biosystems 3730XL DNA sequencer (24). Sequences were listed in supplementary file. Sequences were aligned using ClustalW in MEGA11 with a gap opening penalty of 15.00 and a gap extension penalty of 6.66 (Nehra et al., 2022). Phylogenetic analysis was conducted using the neighbor-joining method and Maximum Likelihood (ML) method with the Tamura-Nei model, employing lowest BIC and AICc values for optimal model selection. Bootstrap analysis with 1,000 replicates was performed to assess statistical support for the phylogenetic tree nodes (Tamura et al., 2013).

## Data Availability

All nucleotide sequences generated in this study are provided in the supplementary file. The authors are in the process of submitting these sequences to a public repository, and the corresponding accession numbers will be updated and made available in the final version of the manuscript upon publication.

## Acknowledgements

The authors greatly acknowledge the National Agricultural Technology Programme (NATP-2), BARC, and Bangladesh for funding (Grant no: NATP-2/PIU-BARC-44/2017-6330, Dated: 25/04/2019). The authors also acknowledge support from Department of Livestock Services, MoFL, Bangladesh; Department of Parasitology, Bangladesh Agricultural University, Mymensingh-2202, Bangladesh. Two abstracts were presented in the 15^th^ International Congress of Parasitology during 21 – 26 August (ICOPA 2022) in Copenhagen, Denmark and 29^th^ International Conference of the World Association for the Advancement of Veterinary Parasitology (WAAVP) during 19-24 August 2023, Chennai, India. The first author, MM Hasan had been awarded a Travel Grant from World Federation of Parasitologists (WFP) in 2022 and the WAAVP Dublin Scholarship in 2023 for his outstanding research.

## Data availability statement

All nucleotide sequences generated in this study are provided in the **supplementary file**. The authors are in the process of submitting these sequences to a public repository, and the corresponding accession numbers will be updated and made available in the final version of the manuscript upon publication.

## Author’s Contribution

MMH and NA: Conceptualization, Methodology, Formal analysis, Resources, Visualization, Writing-Original Draft Preparation, Investigation, Writing-Review & Editing. BCR: Methodology, Draft preparation, Manuscript revision, Formal analysis, Resources, Validation, Software, Writing-Review and Editing, MRR, MMRS, HB and PGB: Methodology, Formal analysis, Writing-Review, A and MZA: Co-supervision, Validation, Visualization, Investigation, Resources, Writing-Review and Editing, MHT: Conceptualization, Methodology, Project Administration, Supervision, Validation, Visualization, Investigation, Resources, Writing-Review and Editing.

